# Sex-Stratified Integrated Analysis of U.S. Lung Cancer Mortality, 1994–2020

**DOI:** 10.64898/2026.03.01.26347234

**Authors:** Muhammad Rafiqul Islam, Sama I. Sayin, Humayera Islam, Mohammad Hasan Shahriar, Muhammad Ashique Haider Chowdhury, Saira Tasmin, Srinivash Konda, Syeda Masuma Siddiqua, Habibul Ahsan

## Abstract

**Importance:** Lung cancer mortality in the United States has fallen substantially in recent decades, yet the relative influence of behavioral, environmental, socioeconomic, and therapeutic factors and their sex specific contributions remains unclear. Understanding these drivers is essential to sustain progress and reduce persistent disparities.

**Objective:** To quantify how behavioral, environmental, socioeconomic, and therapeutic determinants collectively shaped US lung cancer mortality from 1994 to 2020, assess sex specific differences, and forecast mortality trajectories through 2030 using an integrated machine learning framework.

**Design, Setting, and Participants:** Ecological time series study using publicly available national data from 1994 to 2020. Sex stratified analyses were conducted integrating lung cancer mortality, smoking prevalence, fine particulate matter PM2.5 exposure, Human Development Index HDI, per capita healthcare expenditure, healthcare inflation, insurance coverage, income inequality, and annual drug approvals.

**Exposures:** Behavioral smoking, environmental PM2.5, socioeconomic HDI health expenditure inflation, uninsurance inequality, and therapeutic drug approval indicators.

**Main Outcomes and Measures:** Age-standardized lung cancer mortality per 100000 population. Temporal changes were modeled using Joinpoint regression. Concurrent associations were assessed using multivariable and elastic net regression, and forecasts were estimated with AutoRegressive Integrated Moving Average models with exogenous variables ARIMAX.

**Results:** From 1994 to 2020, mortality declined by 59 percent in men, from 52.9 to 21.7 per 100000, and by 40 percent in women, from 26.7 to 15.9 per 100000, with faster declines after 2015. Smoking and PM2.5 decreased by more than 45 percent but remained strongly correlated with mortality. In elastic net models, PM2.5 was the strongest predictor for men, while smoking was the strongest predictor for women. Per capita expenditure and HDI ranked higher for men, while uninsurance and income inequality were strong predictors for women. Mortality declines occurred during periods of major approvals of lung cancer drugs. Forecasts suggest continued but slower declines through 2030, with projected rates of 20.2 and 14.9 deaths per 100000 in men and women, respectively.

**Conclusions and Relevance:** Sex specific declines in lung cancer mortality reflect different dominant correlates, with air pollution more important in men and smoking more important in women, while socioeconomic conditions and therapeutic advances also influence trends. Continued tobacco control, improved air quality, and equitable access to screening and modern treatment are essential to sustain further reductions in mortality.

## Introduction

Lung cancer is the leading cause of cancer-related mortality and the second most commonly diagnosed cancer in the U.S^1^. Since the early 1950s, it emerged as the deadliest cancer for American men, surpassing breast cancer as the leading cause of cancer death among women in 1987^2^. Despite historically high mortality rates, recent epidemiological data reveal a notable decline in lung cancer mortality, averaging a 0.3% annual reduction between 1990 and 2019^3^. This decline marks one of the most significant public health achievements of the early 21st century, which is not attributable to a single breakthrough but rather the result of combined effects of tobacco control^4,5^, cleaner air^6–8^, screening programs^9^, expanded insurance coverage^10^, and diffusion of modern systemic, targeted and precision therapies^11^.

However, mortality declines have not been uniform. Reductions in smoking prevalence align with earlier and larger mortality decreases in men, whereas women show more complex trajectories^12–16^. Heterogeneity likely reflects differences in histological subtype frequencies^17^, historical cohort smoking patterns^18,19^, occupational and outdoor exposures^20^, health-system factors such as screening uptake^21,22^, timeliness of diagnosis^23,24^, biomarker testing^25^, and access to effective therapy within broader social determinants^26^. Long induction periods for lung cancer (20-30 years) further complicate interpretation, because current mortality embeds past exposures while environmental improvements and coverage gains may act on shorter time scales. Importantly, U.S. lung cancer mortality remains higher than in several high-income peers despite rapid recent declines, indicating headroom for further gains through prevention, environmental protection, and equitable access to early detection and effective therapy^27^.

Most prior studies have examined lung cancer mortality in relation to histopathological, behavioral, environmental, socioeconomic, or therapeutic determinants in isolation, limiting insight into their combined population-level influence and how effects vary by sex and over time^5,6,8,11,20,23,25,26,28–31^. To address this gap, we assembled a comprehensive cross-disciplinary dataset curated from multiple major U.S. and international sources^32–42^, and applied an integrated machine-learning based sex-stratified framework evaluating U.S. population-level histopathology, smoking, PM2.5 exposure, Human Development Index (HDI), per capita healthcare expenditure, healthcare expenditure inflation, health insurance coverage, income inequality (Gini coefficient), and lung cancer drug approvals over 1994–2020. We used joinpoint trend analysis, multivariable and penalized regression, elastic net regression and ARIMAX forecasting to estimate sex-specific, concurrent associations across domains and to project near-term trajectories. To our knowledge, this is the first study to integrate detailed datasets across this wide range of variables related to U.S. lung cancer mortality within a single sex-stratified analysis. This integrated assessment evaluates where progress has been greatest, where disparities persist, and which policy levers can most effectively sustain and extend the decline in lung cancer mortality.

## Methods

### Study Design and Data Sources

This ecological, sex-stratified time-series study analyzed national U.S. lung cancer mortality from January 1, 1994, through December 31, 2020. Data were analyzed between January and August 2024. Because only publicly available, de-identified aggregate data were used, this study was exempt from institutional review board approval and informed consent.

Age-standardized lung cancer mortality data were obtained from the World Health Organization (WHO) Cancer Over Time database^42^. Histopathology statistics were collected from the U.S. National Cancer Institute Surveillance, Epidemiology, and End Results (SEER) program^33^. Smoking prevalence data were extracted from the Centers for Disease Control and Prevention (CDC) national surveillance data compiled by the American Lung Association^32^. Annual mean PM2.5 levels were derived from the State of Global Air Initiative dataset^36^. HDI values were obtained from the United Nations Development Reports Portal^39^. Healthcare expenditure inflation data were sourced from the U.S. Inflation Calculator^38^, and per capita healthcare expenditure data were retrieved from the Peterson Center on Healthcare–Kaiser Family Foundation Health System Tracker^35^. Health insurance coverage data were obtained from the U.S. Census Bureau’s Health Insurance Historical Tables (HIB Series)^40^, and income inequality data, expressed as the Gini index, were retrieved from the World Bank development indicators database^41^. To contextualize therapeutic trends, we compiled all U.S. Food and Drug Administration (FDA)–approved lung cancer drugs during the study period from the FDA Drug Database^37^ and the National Cancer Institute (NCI)^34^.

### Variables and Measures

The primary outcome was the age-standardized annual lung cancer mortality rate (per 100 000 population). Independent variables included behavioral (smoking prevalence), environmental (PM2.5 exposure), socioeconomic (HDI, per capita healthcare expenditure, healthcare inflation, uninsurance rate, and Gini index), and therapeutic (annual FDA drug approvals) indicators.

Predictor variables were z-standardized for comparability across models. Scatterplots with fitted regression lines were generated using the seaborn package in Python (version 3.11, Python Software Foundation) to visualize linear associations. Pearson correlation coefficients (r) quantified the strength and direction of bivariate relationships between each predictor and lung cancer mortality, stratified by sex.

### Trend Analysis

Temporal shifts in mortality trends were evaluated using Joinpoint regression to identify statistically significant inflection points. Annual Percent Change (APC) and Estimated Annual Percent Change (EAPC) with corresponding 95% confidence intervals (CIs) were calculated to quantify trend magnitude.

Joinpoints were identified using the Pruned Exact Linear Time (PELT) algorithm in the ruptures package (version 1.1.9) without specifying a fixed maximum number of joinpoints. The algorithm automatically determined the number and location of joinpoints through a penalty-based cost minimization procedure, with a penalty value of 20 to balance model fit and parsimony. Separate models were fit for men and women and, where applicable, by histologic subtype.

### Association Modeling

Annual mortality rates were modeled against behavioral, environmental, and socioeconomic predictors using both linear regression and elastic-net penalized regression. Elastic-net models incorporated both L₁ and L₂ penalties to improve variable selection stability and accommodate collinearity among predictors. Hyperparameters α and λ were optimized via 10-fold cross-validation minimizing mean squared error (MSE). Model performance was evaluated using out-of-sample R² and cross-validated MSE. Regression coefficients were reported as standardized estimates with 95% CIs. Because of long latency between exposure and mortality, smoking coefficients were interpreted as partial correlates rather than causal effects.

### Forecasting Models

To evaluate temporal patterns while accounting for external influences, we applied AutoRegressive Integrated Moving Average models with exogenous variables (ARIMAX), implemented in the statsmodels package (version 0.14). Exogenous predictors included smoking prevalence, PM2.5, and socioeconomic indicators. Model parameters (p, d, q) were selected by the Akaike Information Criterion (AIC) among models satisfying white-noise diagnostics and parameter stability. Model adequacy was confirmed using residual diagnostics, including the Autocorrelation Function (ACF), Partial Autocorrelation Function (PACF), and the Ljung–Box test for independence of residuals.

### Statistical Analysis

Analyses were performed using Python version 3.11 (Python Software Foundation), Joinpoint Regression Program version 5.0.2 (National Cancer Institute), and the statsmodels forecasting library. Two-sided tests were applied with a significance threshold of α = 0.05. For Joinpoint regression, APCs and EAPCs were reported with 95% CIs and permutation test P values. For ridge and elastic-net models, standardized coefficients with 95% CIs were reported alongside cross-validated MSE and out-of-sample R² values.

## Results

### Trends in Lung Cancer Mortality, 1994–2020

Between 1994 and 2020, lung cancer mortality in the United States declined substantially in both sexes, though with distinct trajectories (Figure 1). Among males, the age-standardized mortality rate fell by 59% (from 52.94 to 21.73 per 100 000), while females experienced a 40.3% reduction (from 26.70 to 15.93 per 100 000). Joinpoint regression confirmed these downward trends. For males, the Estimated Annual Percent Change (EAPC) decreased from –2.31% (95% CI, –2.42 to –2.19; P < .001) to –5.62% (95% CI, –6.04 to –5.20; P < .001) (Figure 1B). In contrast, female mortality remained relatively stable between 1994 and 2003 before beginning a significant decline in 2004–2008 (–1.84%; 95% CI, –2.26 to –1.41; P = .0035). The rate of decline accelerated in 2014–2018 (–4.43%; 95% CI, –4.75 to –4.10; P < .001) and remained strong through 2019–2020 (–4.15%; P < .001).

**Figure 1.**
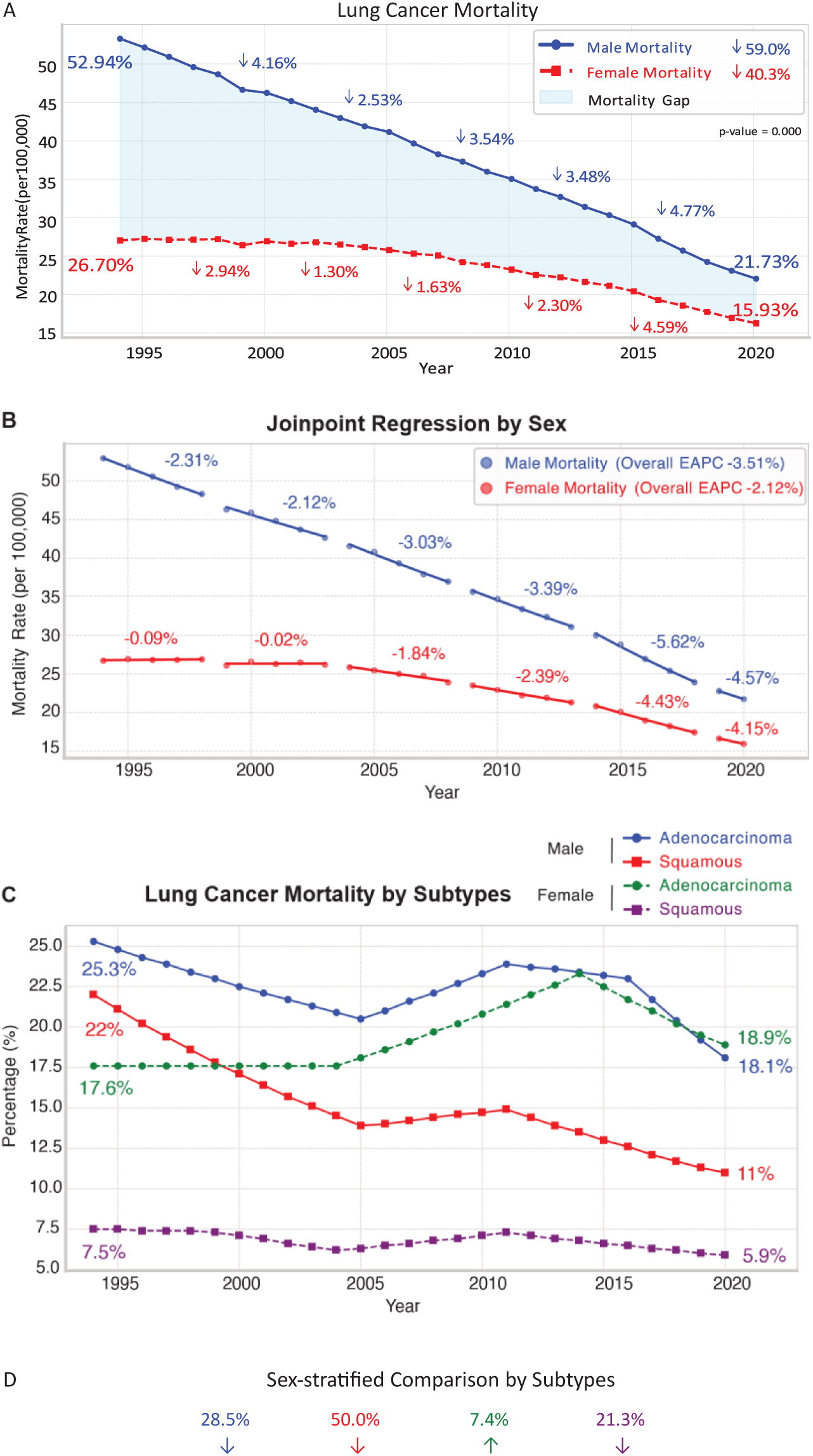
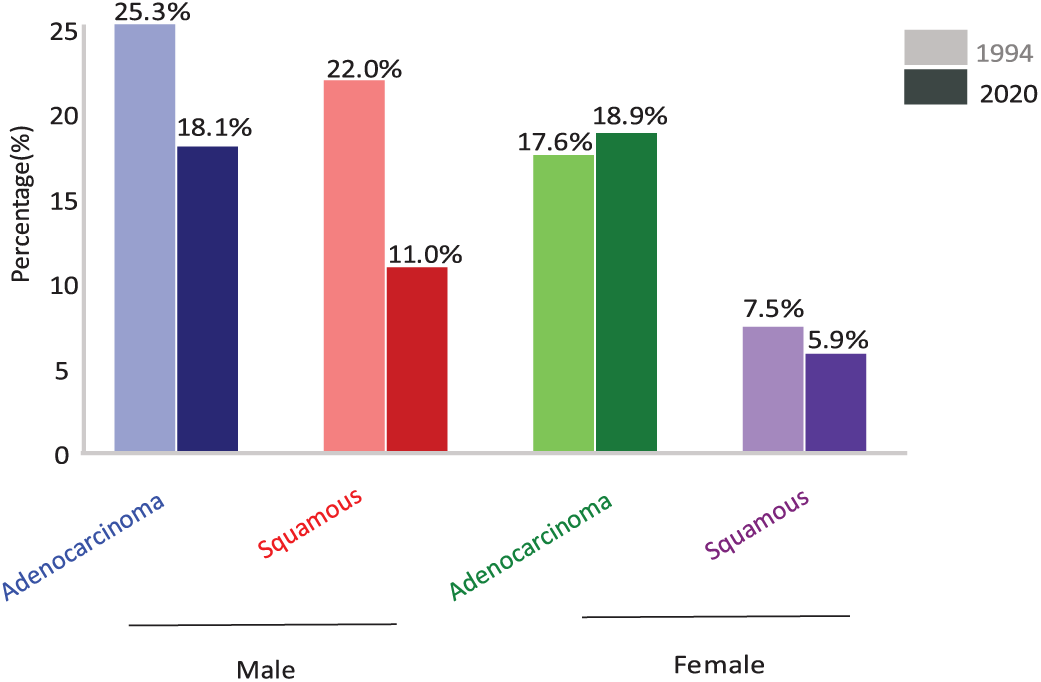
Lung cancer mortality trends in the US (1994-2020). **A,** Overall trends in age-adjusted lung cancer mortality rates stratified by sex. **B,** Joinpoint regression trends stratified by sex. **C,** Lung cancer mortality trends stratified by sex and histologic subtype. **D**, Distribution of adenocarcinoma and squamous cell carcinoma mortality by sex. Abbreviations: EAPC, Estimated annual percent change.

Histologic subtype analyses revealed sex-specific trends. Mortality from squamous cell carcinoma decreased by 50% in men (from 22% to 11%) and by 21.3% in women (from 7.5% to 5.9%) (Figure 1C). Among men, adenocarcinoma mortality declined by 28.5% (from 25.3% to 18.1%), while among women adenocarcinoma initially increased by 32.4% between 2004 and 2014 before declining after 2015, resulting in a net increase of 7.4% (from 17.6% to 18.9%) (Figure 1D). These findings indicate a sustained national decline with earlier and steeper improvement in men and a delayed downturn in women, partly driven by differing adenocarcinoma trajectories.

### Smoking and PM2.5

Smoking prevalence declined steadily from 27.0% to 14.1% (47.8% reduction) in men and from 22.6% to 11.0% (51.3% reduction) in women, yet mortality remained strongly correlated (r = 0.98 for men, r = 0.96 for women) (Figure 2A). Linear regression showed that each 1% increase in smoking prevalence was associated with a 2.14-fold increase (95% CI,1.98 to 2.32; P < .001) in male mortality and a 0.96-fold increase in female mortality (95% CI, 0.85 to1.12; P < .001).

**Figure 2.**
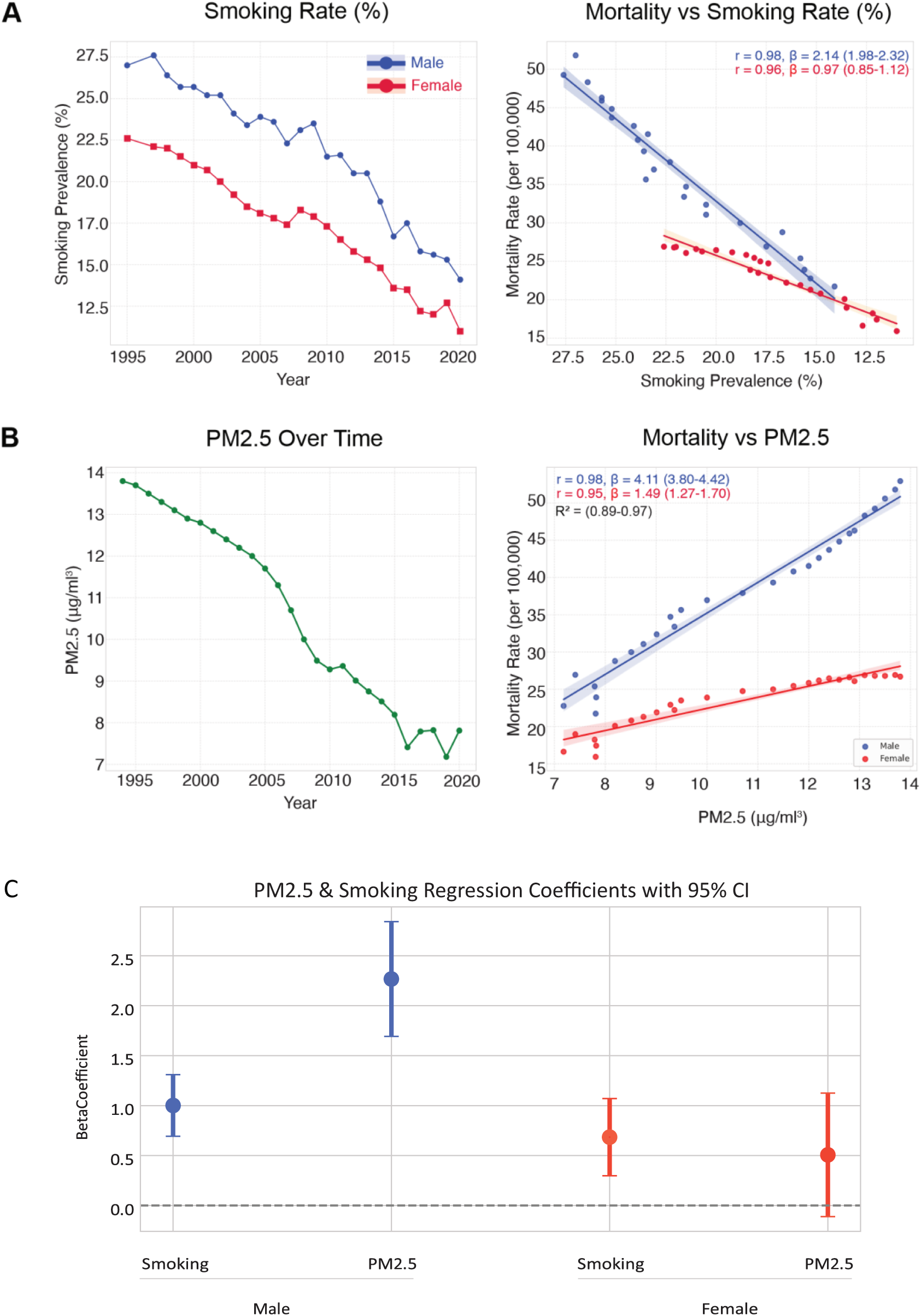
Lung cancer mortality in the US in relation to smoking and PM2.5. Temporal trends of **A**, smoking rates (left) and **B**, PM2.5 (left) and corresponding Pearson correlation (right) with male (red) and female (blue) lung cancer mortality. **C**, Smoking-adjusted regression coefficients with 95% CI (error bars). Abbreviations: PM2.5, Fine particulate matter. r, Pearson correlation. β, Linear regression beta coefficient

Ambient PM2.5 levels declined from 13.8 μg/m³ in 1994 to 7.81 μg/m³ in 2020, with sharpest declines coinciding with the sharpest mortality reduction between 2004 and 2009 (20.9%) (Figure 2B). Regression analysis revealed a stronger association between PM2.5 and male mortality (β = 4.11; 95% CI, 3.80 to 4.42; P < .001) than female mortality (β = 1.49; 95% CI, 1.27 to 1.70; P < .001), with R² values ranging from 0.89 to 0.97. After adjustment for smoking, each 1 μg/m³ increase in PM2.5 corresponded to an additional 2.26 male deaths per 100 000 (95% CI, 1.69 to 2.84; P < .001), while the association in females was weaker and not statistically significant (0.50 per 100 000; 95% CI, –0.11 to 1.12; P = .10) (Figure 2C).

### Socioeconomic Indicators

Between 1994 and 2019, HDI rose from 0.885 to 0.933, while healthcare inflation declined from 4.78% to 1.98% (Figure 3A). HDI was strongly inversely correlated with mortality (r = –0.98 for men; r = –0.92 for women), whereas healthcare inflation showed a modest positive correlation (r = 0.54 for men; r = 0.56 for women) (Figure 3B).

**Figure 3.**
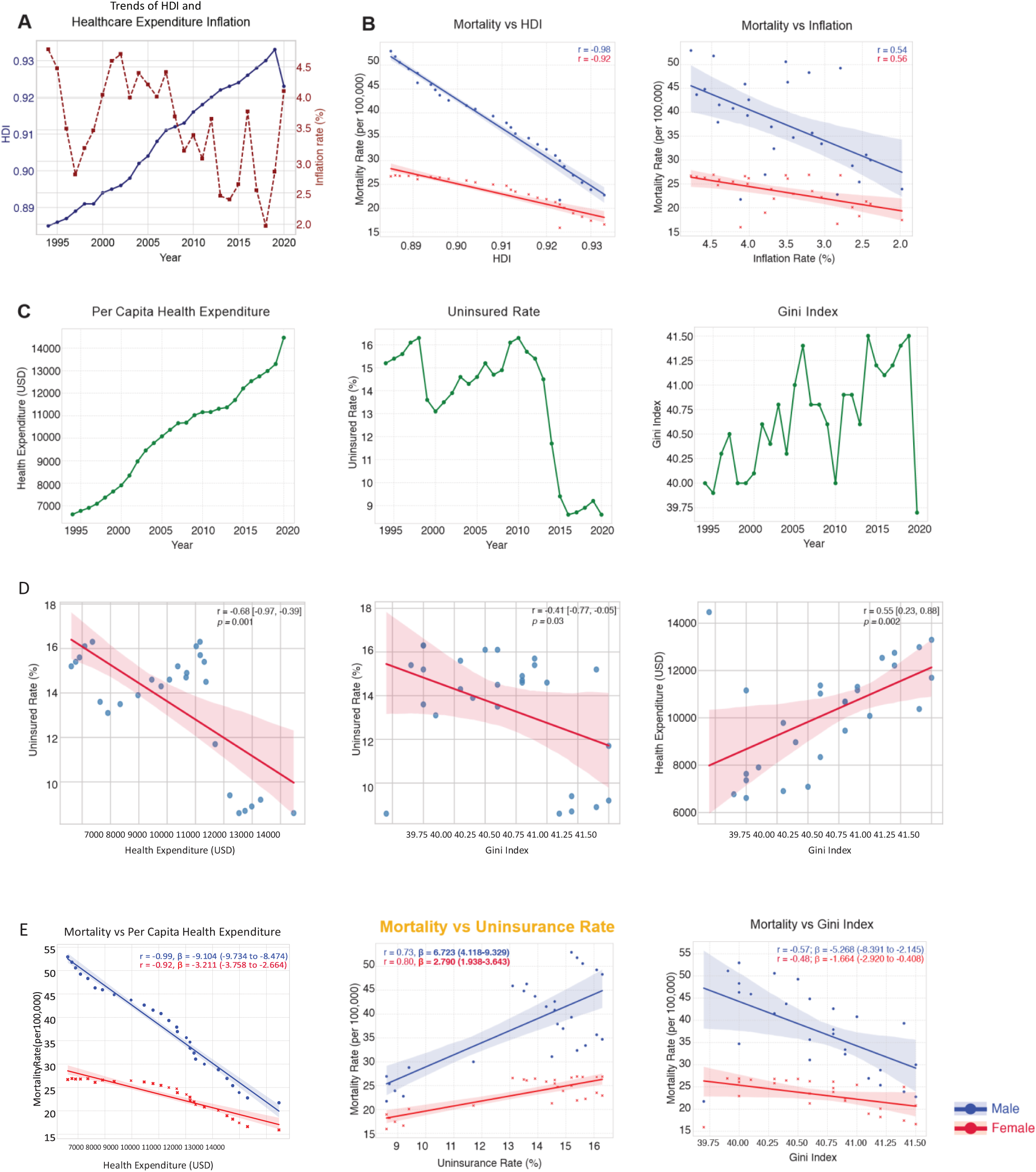
Lung cancer mortality in the US in relation to socioeconomic determinants. A, Trends of Human Development Index (HDI) and healthcare expenditure inflation over time (left) and corresponding standardized regression analysis (right). B, HDI (left) and inflation (right) in relation to mortality rates in males (blue) and females (red). C, Trends of per capita health expenditure (left), uninsurance rate (middle) and Gini index (right) over time. D, Standardized regression analysis of interaction between variables in C. E, Per capita health expenditure (left), uninsurance rate (middle) and Gini index (right) in relation to mortality rates. r, Pearson correlation. β, Linear regression beta coefficient

Per capita healthcare expenditure more than doubled (from $6616 to $14 466) over the same period, the uninsured population declined from 15.2% to 8.6%, and the Gini index increased modestly (from 39.7 to 41.5) (Figure 3C). Inverse correlations were observed between the uninsured rate and both healthcare expenditure (r = –0.68; 95% CI, –0.97 to –0.39; P = .0001) and the Gini index (r = –0.41; 95% CI, –0.77 to –0.05; P = .03); however, the Gini index and per capita expenditure were positively associated (r = 0.55; 95% CI, 0.23 to 0.88; P = 0.002) (Figure 3D). The uninsured rate correlated positively with mortality (r = 0.73 for men; r = 0.80 for women), whereas healthcare expenditure correlated inversely (r = –0.99 for men; r = –0.92 for women). The Gini index demonstrated weaker negative correlations (r = –0.57 for men; r = –0.48 for women) (Figure 3E).

Standardized regression analyses further revealed that per capita healthcare spending was inversely related to mortality in males (β = –9.10) and females (β = –3.21), but the uninsured rate showed a positive association with mortality (β = 6.72 for males, 2.79 for females). The Gini index was negatively associated with lung cancer mortality for both sexes (β = –5.26 for males, –1.66 for females) (Figure 3E).

### Therapeutic Approvals and Temporal Alignment

During the study period, the FDA approved 30 new lung cancer drugs, including 16 targeted therapies (mostly tyrosine kinase inhibitors), 8 immunotherapies, and 6 chemotherapies (Figure 4A). Periods of accelerated mortality decline aligned temporally with these approval waves. Among men, the steepest decline (2014–2017) corresponded to a reduction of 6.08 per 100 000, while among women the steepest decline (2016–2019) was 3.47 per 100 000 (Figure 4B). Annual drug approvals correlated inversely with mortality (r = –0.51 for men; P = .0065 and r = –0.56 for women; P = .0024).

**Figure 4.**
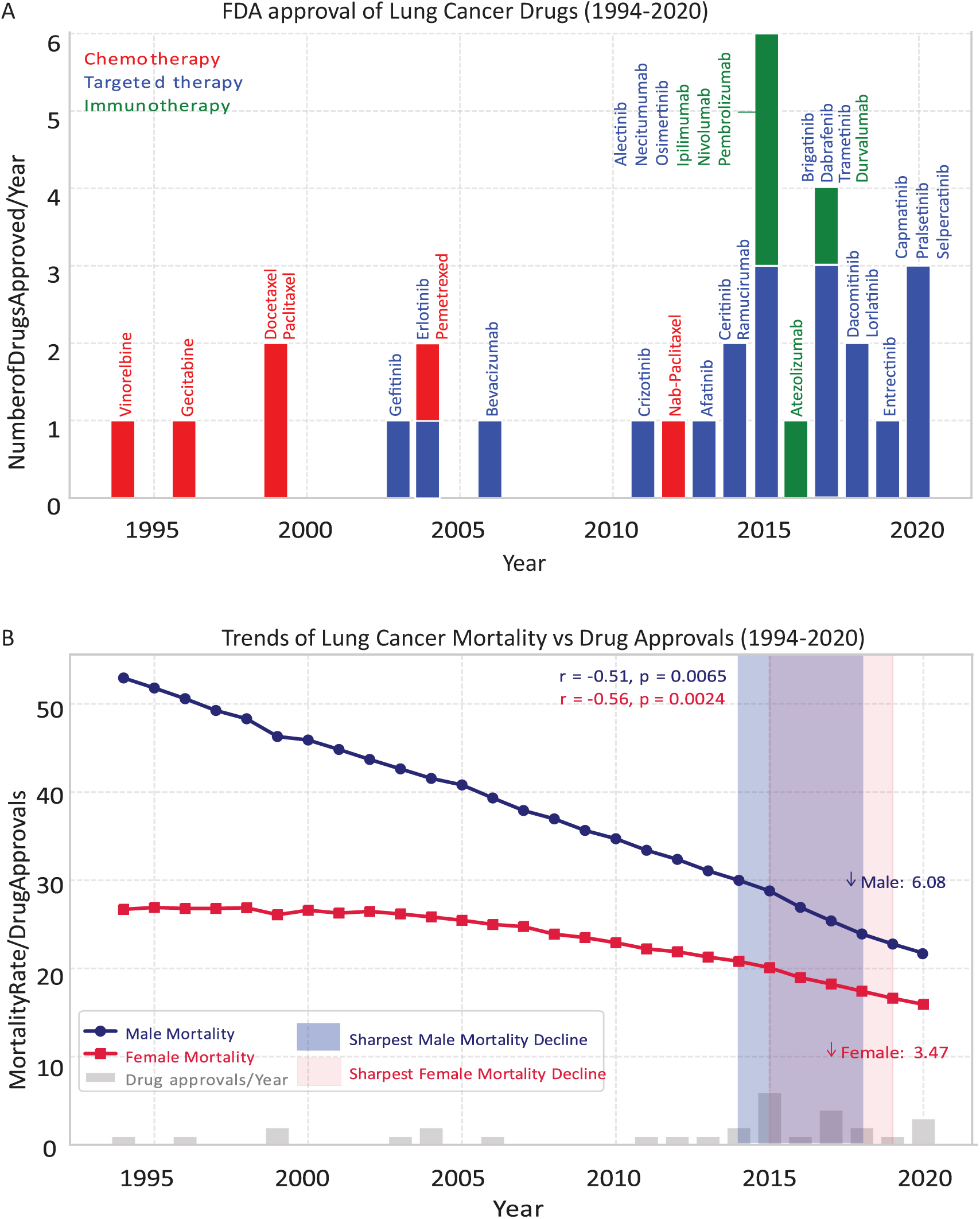
Lung cancer mortality in the US in relation to drug approval trends. **A**, Annual count of lung cancer drugs approved by the FDA (1994-2000), categorized by type: chemotherapy (red), targeted therapies (blue), and immunotherapies (green). **B**, overlays the regression-based trends in lung cancer mortality for males (blue) and females (red) with the periods during which the major drug types were introduced. Shaded vertical regions indicate the eras with sharpest fall in mortality for males (lavender) and females (light red). r, Pearson correlation.

### Integrated Predictors and Machine Learning Modelling

Elastic-net regression models identified distinct sex-specific patterns in predictor importance (Figure 5A). Among men, PM2.5 had the largest standardized coefficient, followed by per capita expenditure and HDI, while smoking ranked fourth. Among women, smoking prevalence had the strongest positive coefficient, followed by uninsurance and income inequality. HDI showed opposite effects by sex – protective in men but slightly positive in women. Healthcare inflation ranked low in both sexes, indicating a modest role after adjustment for other variables.

**Figure 5.**
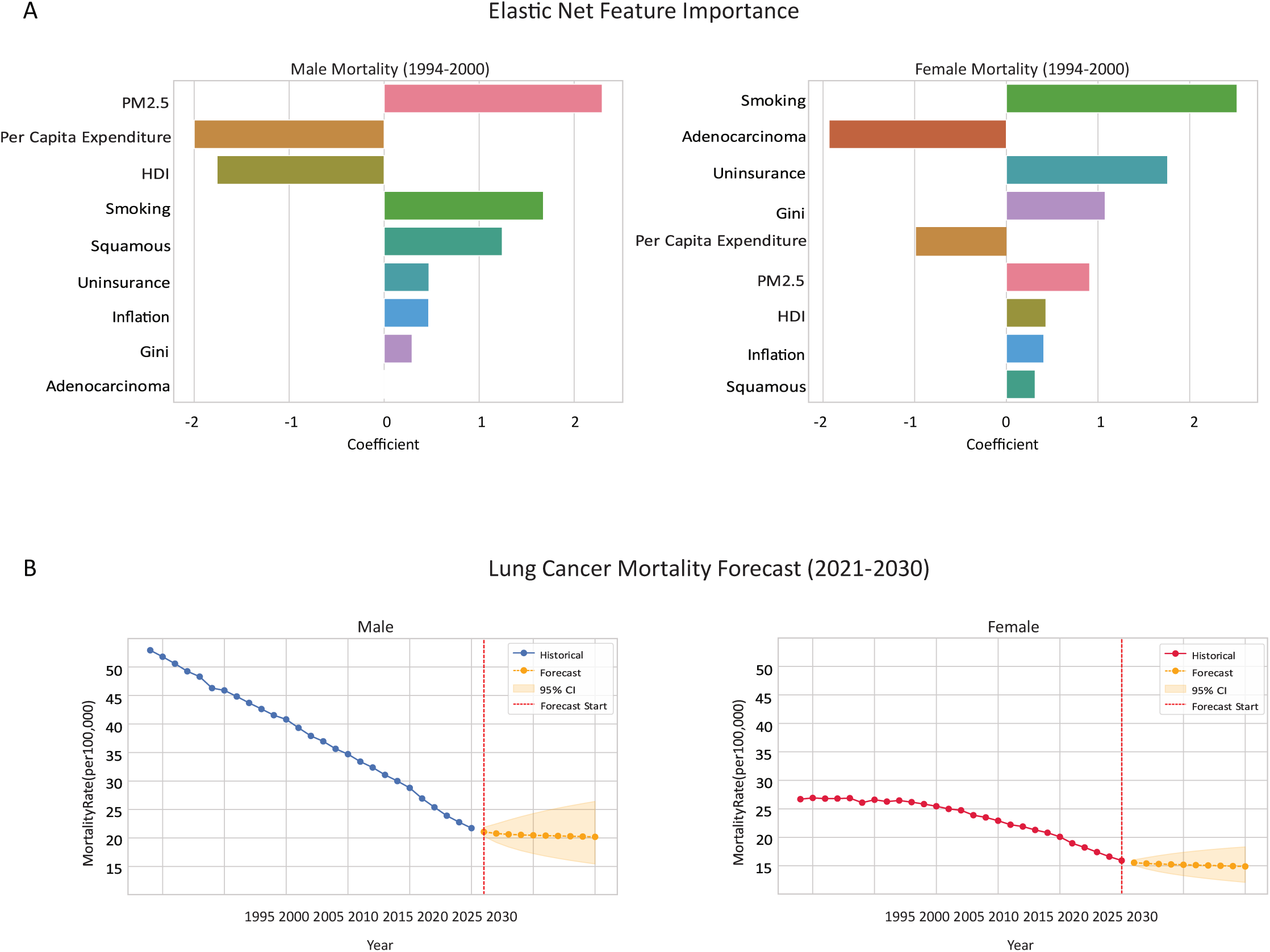
Integrated analysis of behavioral, environmental and socioeconomic determinants of lung cancer mortality in the US. **A**, Elastic Net feature importance for male (left) and female (right) lung cancer mortality, displayed as standardized coefficients. **B**, Forecasted lung cancer mortality rates in the United States for males (left) and females (right) using ARIMAX modeling (2021–2030). ARIMAX models were applied on log-transformed mortality data using smoking prevalence as an external regressor. The solid blue lines represent historical mortality data (1994–2020), and the dashed orange lines depict forecasts for 2021–2030. Shaded areas represent 95% confidence intervals for the forecasted estimates. A vertical red dashed line indicates the start of the forecast period in 2021.

### Forecasts of Lung Cancer Mortality Through 2030

ARIMAX models incorporating external predictors (smoking, PM2.5, socioeconomic indicators) projected a continued but slower decline in lung cancer mortality through 2030 (Figure 5B). Male mortality is forecast to decrease from 21.1 to 20.2 deaths per 100 000 (95% CI, 15.4 to 26.4), and female mortality from 15.6 to 14.9 deaths per 100 000 (95% CI, 12.1 to 18.3). Forecast uncertainty increased with projection length, especially among women, as indicated by widening 95% CIs.

## Discussion

Lung cancer mortality in the United States has declined sharply since 1994, but men and women have not shared this progress equally. Using an integrated, sex-stratified framework that incorporated behavioral, environmental, socioeconomic, and therapeutic factors, we clarified these differences and identified key determinants. The dominant correlates of mortality diverged by sex – PM2.5 for men and smoking for women – while socioeconomic and insurance effects weakened when behavioral and environmental variables were modeled together. Mortality trends also aligned with therapeutic approval waves, suggesting sex-specific diffusion of treatment benefit. Considering these domains together strengthens interpretation of national patterns and points to practical, sex-aware levers for prevention and care.

Men and women traversed the mortality decline on distinct timelines, and the present dataset quantifies those differences with greater overview and temporal resolution than most prior studies^12–14,43^. Nationally, male mortality declined nearly 60% compared with 40% in females, with Joinpoint analysis revealing earlier and steeper turning points among men. These trajectories align with earlier studies attributing slower female declines to later smoking peaks and adenocarcinoma predominance^14,18^. Extending observation through 2020 reveals rapid narrowing of the mortality gap after 2015, driven by accelerated declines in female adenocarcinoma mortality that to our knowledge has not been reported previously. The United States now approaches convergence in sex-specific mortality, enabling finer interpretation of behavioral and socioeconomic drivers.

Joint modeling confirmed different leading predictors by sex: PM2.5 in men and smoking in women. This reflects exposure magnitude and timing: female mortality still tracks smoking lags of 20–30 years, whereas men experience greater occupational and outdoor particle exposure^18^. The national PM2.5 decline since the mid-2000s parallels the earlier male mortality drop^7,8^. Fine particles reach the peripheral lung, promoting adenocarcinoma and EGFR-mutant epithelial clones, an effect that is not sex-specific ^6,8^. Although PM2.5 likely increases risk in both sexes, women’s higher prevalence of targetable mutations confers better survival, muting its effect on mortality. Thus, ambient pollution explains more residual variation in men, while smoking remains dominant in women.

Socioeconomic and insurance variables contributed modestly once behavioral and environmental factors were included. In men, per-capita income and HDI were retained with negative coefficients; in women, uninsurance and Gini ranked higher. These results align with prior evidence that coverage improves timely diagnosis and treatment access. ^10,26^, The instability of inequality measures after adjustment likely reflects concurrent improvements in coverage and spending rather than any protective role for inequality.

Therapeutic approvals provided additional context. The steepest mortality declines coincided with rising annual drug approvals (2014–2017 in men and 2016–2019 in women), consistent with evidence that modern systemic therapies improve survival, even though prevention remains the main driver of population benefit ^11,29^.

Elastic Net modelling suggested that mortality trends in men are most influenced by environmental exposure (PM2.5) and structural socioeconomic factors (per capita expenditure, HDI), whereas in women behavioral (smoking) and access-related variables (uninsurance and income inequality) predominate. Forecasts extend these patterns. The projections are of continued but slowing declines through 2030 with a persistent, narrower sex gap. Interpreted with the coefficient profiles, the implication is that marginal gains will increasingly depend on targeting the dominant correlates in each sex while sustaining the system capacity that enables equitable access, prevention, early detection, and effective treatment.

### Limitations

This study has limitations. First, as an ecological, population-level analysis, the associations identified between behavioral, environmental, socioeconomic, and therapeutic factors and lung cancer mortality may not represent causal relationships at the individual level. Second, racial, geographic, and age-specific differences could not be assessed because national aggregate data were used. Third, temporal alignment of exposures and outcomes is limited by the long induction period of lung cancer, meaning that current mortality partly reflects exposures from decades earlier. Fourth, the use of FDA drug approvals as a proxy for therapeutic diffusion does not capture clinical utilization, treatment adherence, or survival benefit at the patient level. Finally, the analyses relied on publicly available datasets that may differ in completeness and granularity across time, potentially introducing measurement bias. Despite these limitations, the integrated, sex-stratified, and machine-learning–based approach strengthens inference by combining otherwise widespread domains within a unified analytic framework.

## Conclusion

In this national, sex-stratified analysis integrating behavioral, environmental, socioeconomic, and therapeutic determinants, U.S. lung cancer mortality declined sharply from 1994 to 2020 but through distinct pathways in men and women. Air pollution was the dominant correlate among men, whereas smoking remained strongest among women, and socioeconomic factors and therapeutic advances contributed secondary effects. Structural socioeconomic factors influenced mortality more among men, whereas in women access-related variables dominated. These findings highlight the importance of sustaining tobacco control, enforcing clean air standards, and ensuring equitable access to early detection and modern therapy. Continued progress will depend on integrating prevention, environmental policy, and health system equity to close remaining gaps in lung cancer outcomes.

## Data Availability

All data produced in the present study are available upon reasonable request to the authors

## Article Information

## Author Contributions

Dr Islam had full access to all the data in the study and takes responsibility for the integrity of the data and the accuracy of the analysis.

Concept and design: Islam, Sayin.

Acquisition, analysis, or interpretation of data: Islam, Sayin, Shahriar, Chowdhury, Tasmin, Konda, Siddiqua, H. Islam.

Drafting of the manuscript: Islam, Sayin.

Critical revision of the manuscript for important intellectual content: All authors.

Statistical analysis: Islam, Konda.

Administrative, technical, or material support: Shahriar, Chowdhury, Tasmin, Siddiqua.

Supervision: Ahsan.

## Data Sharing Statement

All analyses used publicly available, de-identified aggregate data and therefore did not constitute human subject research. Source datasets are specified in respective reference. Analytical codes will be made available upon reasonable request to the corresponding author.

## Acknowledgments

We thank the national and international public data repositories whose open datasets made this analysis possible.

